# Algorithmic Individual Fairness and Healthcare: A Scoping Review

**DOI:** 10.1101/2024.03.25.24304853

**Authors:** Joshua W. Anderson, Shyam Visweswaran

## Abstract

**Objective:** Statistical and artificial intelligence algorithms are increasingly being developed for use in healthcare. These algorithms may reflect biases that magnify disparities in clinical care, and there is a growing need for understanding how algorithmic biases can be mitigated in pursuit of algorithmic fairness. Individual fairness in algorithms constrains algorithms to the notion that “similar individuals should be treated similarly.” We conducted a scoping review on algorithmic individual fairness to understand the current state of research in the metrics and methods developed to achieve individual fairness and its applications in healthcare.

**Methods:** We searched three databases, PubMed, ACM Digital Library, and IEEE Xplore, for algorithmic individual fairness metrics, algorithmic bias mitigation, and healthcare applications. Our search was restricted to articles published between January 2013 and September 2023. We identified 1,886 articles through database searches and manually identified one article from which we included 30 articles in the review. Data from the selected articles were extracted, and the findings were synthesized.

**Results:** Based on the 30 articles in the review, we identified several themes, including philosophical underpinnings of fairness, individual fairness metrics, mitigation methods for achieving individual fairness, implications of achieving individual fairness on group fairness and vice versa, fairness metrics that combined individual fairness and group fairness, software for measuring and optimizing individual fairness, and applications of individual fairness in healthcare.

**Conclusion:** While there has been significant work on algorithmic individual fairness in recent years, the definition, use, and study of individual fairness remain in their infancy, especially in healthcare. Future research is needed to apply and evaluate individual fairness in healthcare comprehensively.

## 1 Introduction

Statistical and artificial intelligence (AI) algorithms^†^ have improved clinicians’ ability to provide quality healthcare. Such algorithms have accelerated healthcare discoveries, improved clinical decision-making, and lowered costs in healthcare.^15^ However, ethical concerns have been raised about the potential for such algorithms to exacerbate already-existing disparities among marginalized populations.^29^

Algorithmic fairness in healthcare is critical for ensuring equitable assessment and treatment of all individuals, regardless of their background. Various biases can creep into algorithmic development and application, affecting the fairness of such algorithms.^9^ A range of protected attributes, factors that should not influence health, have been chosen because of legal mandates or because of organizational values.^65^ Some common protected attributes include race, ethnicity, religion, national origin, gender, marital status, age, and socioeconomic status. Yet, several healthcare algorithms have been shown to be unfair, particularly across racial categories,^64^ and nearly 50 clinical algorithms are in use that include race, a key protected attribute, as an input variable.^63^

### 1.1 Unfairness in healthcare algorithms

Broadly speaking, biases in statistical and AI algorithms are caused by three factors: i) unrepresentative data used for algorithm development (data bias), ii) poor design in algorithm development (development bias), and iii) improper user - clinician or patient - interactions with the algorithm (interaction bias).^52^ Biases in data are problems that arise from a variety of issues related to data collection and organization and include minority bias, missing data bias, informativeness bias, and selection bias.^60^ Minority bias occurs when there are insufficient data from minority groups to develop an accurate algorithm (e.g., the data includes far too few members of racial minority groups^40^). Missing data bias occurs when data from minority groups are missing systematically making it difficult to learn accurate statistical patterns (e.g., members of racial minorities with limited access to healthcare have fewer electronic health record data^30^). Informativeness bias occurs when data and features used by an algorithm are less useful in a minority group (e.g., detecting melanoma in patients with dark skin is more difficult than in those with light skin^1^). Selection bias occurs when the data used to develop an algorithm is not representative of the population it will be deployed (e.g., data from a single healthcare system may not be representative of other healthcare systems^68^). Observational data such as from electronic health records (EHRs) that are increasingly used in developing algorithms, likely introduce more biases than carefully curated data from research studies, due to inadequate documentation, ambiguous or varying definitions, and other systematic issues.

Despite utilizing unbiased and representative data, algorithms may still manifest bias due to poor design in algorithm development. Examples of such development bias issues encompass label bias and cohort bias.^52^ Label bias occurs when algorithm development employs inconsistent labels, which do not mean the same thing for all individuals because they are an imperfect proxy. For example, racial bias was identified in an algorithm that predicted future healthcare needs of patients because the data that was used in development employed medical cost as a surrogate for healthcare utilization.^47^ Cohort bias arises when the development of an algorithm is predicated on conventional or readily quantifiable groups, neglecting to consider alternative protected groups or granularity levels. For example, mental health disorders have been underdiagnosed within lesbian and gay groups due to algorithms that fail to account for the granularity of sex and instead rely solely on the binary recording of male or female.^45^

Interaction biases can occur when healthcare providers or patients interact with algorithms in ways that affect the algorithm’s performance and fairness.^60^ Automation bias is an example of clinician-interaction bias in which clinicians are unaware that an algorithm is less accurate for a specific group and place too much trust in it, accepting incorrect recommendations.^5^ Privilege bias is a type of patient-interaction bias that occurs when algorithms are not available in settings where protected groups receive care, resulting in unequal distribution of algorithmic healthcare benefits.^24^

### 1.2 Measuring algorithmic fairness

To characterize algorithmic fairness, measures to assess fairness or, equivalently, bias are needed. Broadly speaking, two types of fairness metrics have been described: group and individual notions of fairness.^20^

Most of the literature focuses on the first notion of fairness, which is based on parity of statistical metrics across groups that differ in a protected attribute (e.g., male and female groups). A wide range of group fairness (GF) metrics have been developed and described in the literature. Popular measures include parity of positive prediction rate (known as demographic parity^22^), parity of true positive rates (known as equal opportunity^32^), parity of false positive and false negative rates (known as equalized odds^32^), and parity of positive predictive value^19^ across protected groups. Comprehensive lists of GF metrics are described in the literature.^4, 12^ These metrics are appealing because they are simple to define and measure, and they are based on well-known performance metrics used for algorithmic evaluation. However, statistical definitions of fairness do not give meaningful guarantees to individuals; rather, they provide guarantees on average for groups. Furthermore, it is impossible to optimize multiple GF metrics simultaneously. For example, except in trivial settings, it is impossible to equalize false positive rates, false negative rates, and positive predictive value across groups simultaneously.^39^ One significant disadvantage of enforcing GF metrics is that, because they only require parity to be satisfied to the level of groups, bias can occur within groups at the individual or subgroup levels. Individual notions of fairness, on the other hand, guarantee that similar individuals receive the same decisions from algorithms. It focuses on ensuring fairness for each individual, regardless of their group membership. This means that individuals with similar relevant characteristics (e.g., comorbidities) should be treated similarly by the algorithm regardless of protected characteristics such as race or sex. Compared to group fairness, individual fairness (IF) is less frequently described in the literature. Dwork et al. was the first to propose that “similar individuals should be treated similarly,” with similarity between pairs of individuals defined in terms of a task-specific metric.^22^ According to Joseph et al., “less qualified individuals should not be favored over more qualified individuals,” where quality is defined with respect to the true underlying label that the algorithm does not know.^36^ Kusner et al. proposed a type of IF called counterfactual fairness.^41^ Counterfactual fairness (CF) is a principle for ensuring fairness that states that a decision is fair if it would be the same for an individual even if their protected attributes (e.g., race, gender) were different in a counterfactual world. This means the algorithm’s decision is not affected by group membership, but only by their relevant characteristics of an individual.

It is worth noting that the term fairness is frequently used in the literature to refer to two distinct concepts: a metric that measures the fairness of an algorithm and a constraint applied while developing a fair algorithm. For example, the concept of “equal opportunity” in GF refers to a fairness metric, defined as the difference of the true positive rates between two groups, and a fairness constraint, expressed as a difference of the true positive rates close to zero.^48^ Often, the context in which the term fairness is used will clarify whether it refers to a metric or a constraint.

### 1.3 Motivation

Our examination of IF was prompted in part by a rough parallel in the domain of predictive modeling. Statistical and AI approaches for training predictive algorithms can be broadly categorized into population-wide and patient-specific modeling that have rough parallels to GF and IF. The conventional predictive modeling approach in healthcare (and other areas) consists of learning a single algorithm from a database of individuals, which is then applied to decisions for each future individual. Such a model is called a population-wide model since it is intended to be applied to an entire population of future individuals and is optimized to have good predictive performance on average on all members of the population.^61^ Patient-specific modeling, on the other hand, focuses on learning models that are tuned to the characteristics of the individual at hand, and such models are optimized to perform well for a specific individual.^61^ Many patient-specific methods depend on assessing the similarity between individuals and hence use a similarity method. The canonical technique is the k-nearest neighbor method, which predicts the outcome in an individual based on a group of k-nearest individuals in the data. Other patient-specific methods train a model that is influenced by the characteristics of the patient at hand without using a similarity measure.^35, 42, 61, 62^

While Dwork et al., Joseph et al., and Kusner et al. provide notions of IF, ambiguity and heterogeneity persist in the domain of IF. Furthermore, we discovered no existing literature reviews on IF, which was the primary motivation for conducting this review.

## 2 Methods

We determined that a scoping review was appropriate due to the lack of existing literature reviews on this topic, our desire to broadly summarize the approaches to IF, and the potential role of IF in mitigating algorithmic bias. We followed the methodological framework of Arksey and O’Malley and the Preferred Reporting Items for Systematic review and Meta-Analyses extension for Scoping Reviews.^6, 59^ We performed this scoping review in the following steps: (1) identify the research questions, (2) find relevant articles, (3) select articles, (3) extract data and themes, and (5) report the findings. We describe the first four steps here and report the findings in the “Results” section.

### 2.1 Identifying the research questions

The purpose of this study was to conduct a scoping review of the literature on IF to describe current approaches to IF and explore the potential role of IF methods in mitigating algorithmic bias. IF methods relevant to this review were defined according to the descriptions presented in Dwork et al., “similar individuals being treated similarly,” Joseph et al. “less qualified individuals should not be favored over more qualified individuals,” and in Kusner et al., who presented that individual decisions should “remain unchanged in a world where an individual’s protected attributes had been different in a causal sense.”^22, 36, 41^ Specifically, the review was conducted to address the gap in understanding of the characteristics of IF methods and their scope of use by addressing the following two research questions:

1. What notions of similarity are used in IF?
2. How is IF used in mitigating bias in algorithms?

### 2.2 Finding relevant articles

We searched for relevant articles and conference proceedings in three databases: PubMed, ACM Digital Library (DL), and IEEE Xplore. Because we wanted to retrieve as many relevant articles as possible, we devised a search strategy that prioritized recall over precision. Since the term “fairness” spans many disciplines in forms that are not algorithmic, we developed distinct search queries for each database to adjust for their relative sensitivities concerning these non-algorithmic notions of fairness. The search query for PubMed included the term “algorithmic individual fairness” appearing in the title or abstract. The search query for ACM DL included the term “algorithmic individual fairness” appearing in the title or abstract along with one or more of the following terms: “fair AI”, “individual model fairness.” The search query for IEEE Xplore included the term “individual fairness” appearing in the title or abstract along with one or more of the following terms: “algorithmic”, “model”, “machine learning”, or “artificial intelligence.” The database-specific fields and queries we used are summarized in Table 1.

**Table 1:**
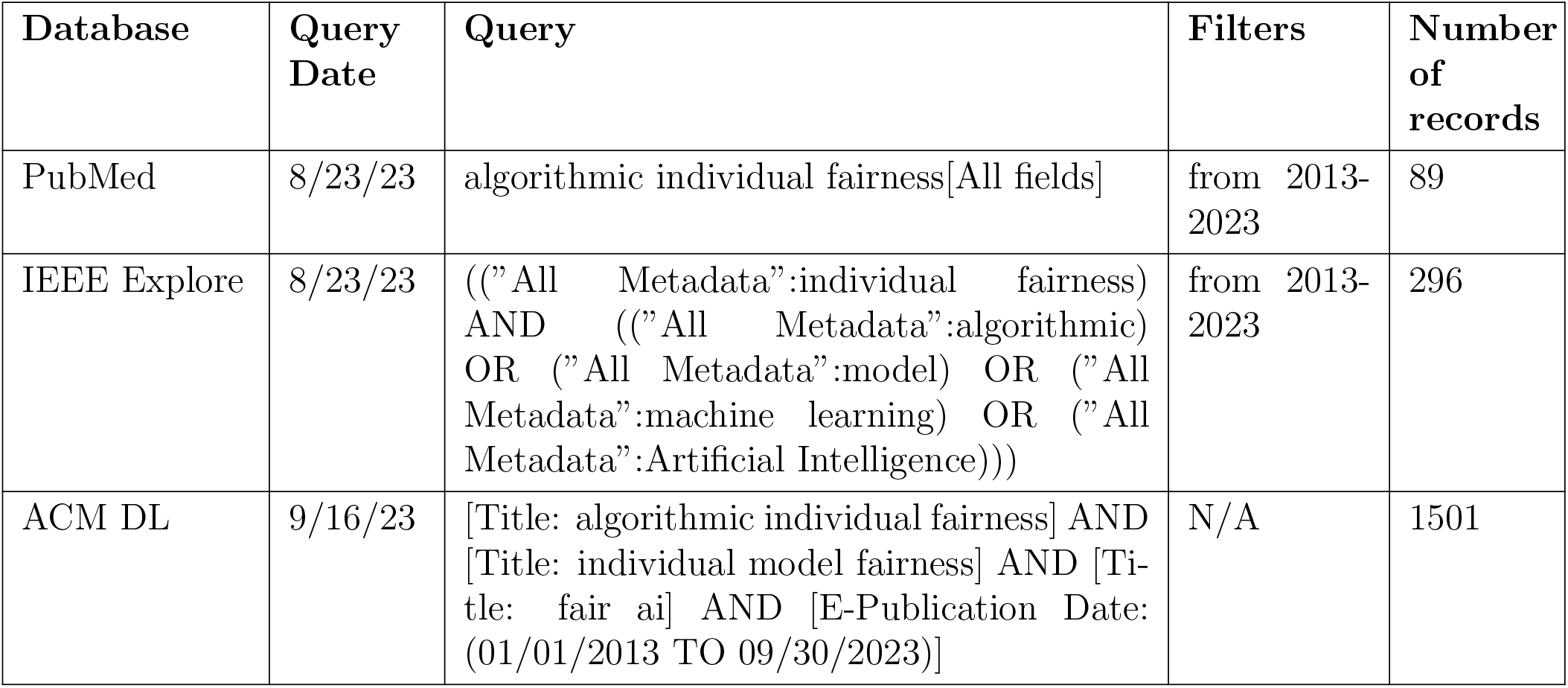
Database queries for identifying relevant articles.

### 2.3 Selection of articles

We reviewed the titles and abstracts of unique articles obtained in the first step to identify articles for a full-text review (see Figure 1). We selected articles that studied IF methods and their uses based on the following inclusion and exclusion criteria. We included articles that described the following:

**Figure 1:**
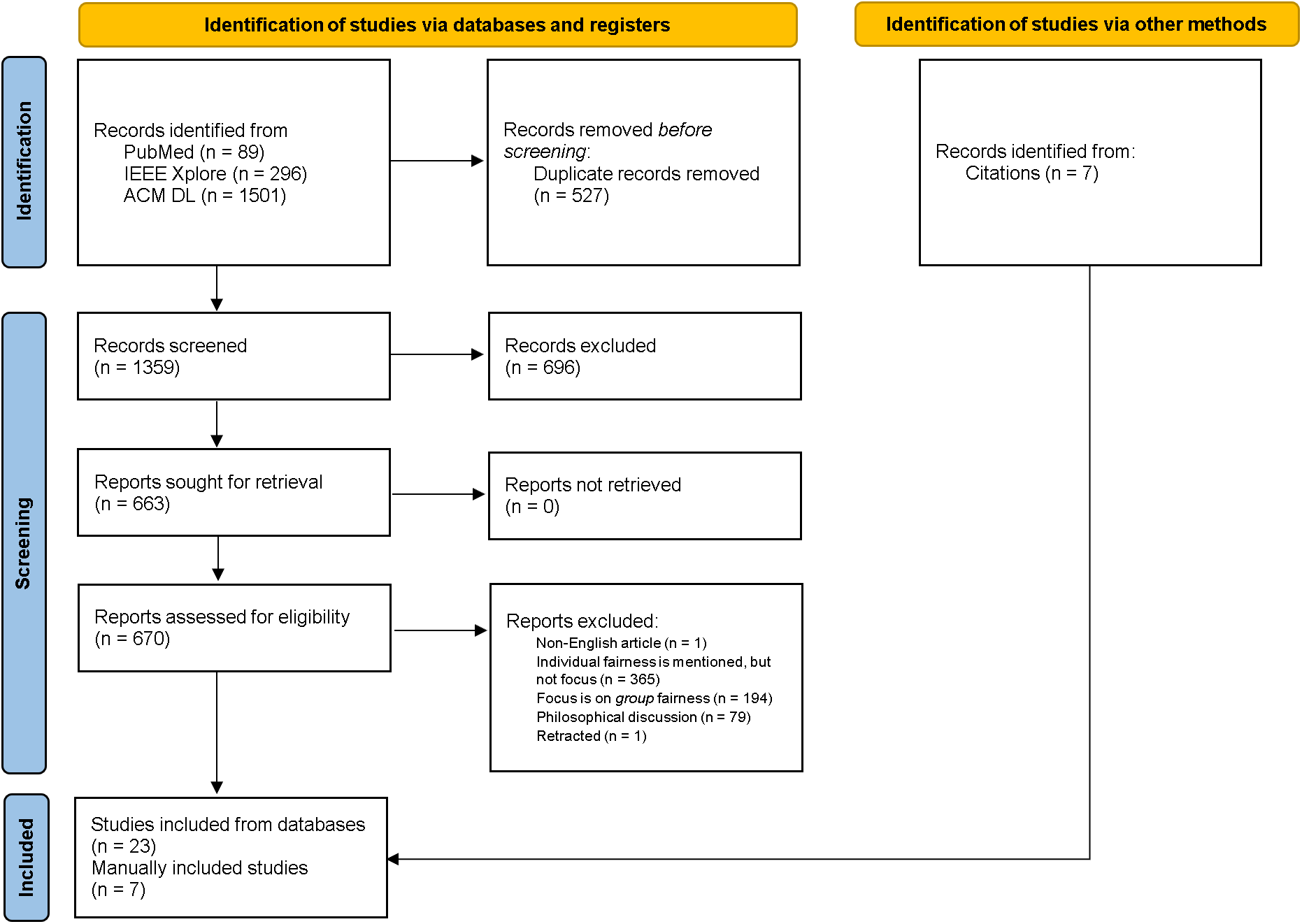
Preferred Reporting Items for Systematic Reviews and Meta-Analyses (PRISMA) diagram of the article screening process.

- studies that focused on algorithmic IF
- studies that compared IF with GF

We excluded articles that described the following:

- mentioned or briefly described algorithmic IF, but IF was not the focus
- studies that focused on algorithmic GF
- exclusively philosophical discussions of fairness including algorithmic fairness
- not written in the English language
- articles that were later retracted

Because of the broad search criteria, many of the articles returned were not specifically about algorithmic IF. A number of articles that were found discussed differential privacy. Differential privacy and algorithmic fairness are closely related, and methods from differential privacy have been used to develop notions of algorithmic IF.^22^ Despite this connection, we decided to leave these articles out of the final list because we preferred explicit notions of algorithmic IF. We identified a group of relevant articles for the review using the inclusion and exclusion criteria. Reasons for exclusion were recorded for the excluded articles. The PRISMA-ScR flow diagram displays the number of excluded papers as well as the reasons for exclusion (see Figure 1)

### 2.4 Data extraction

We extracted information from articles and entered it into a spreadsheet for analysis. We recorded the year of publication, the similarity metric or methodology used, fairness mitigation methods, and the notion of IF for each article. In addition, we included a summary of each article’s findings. The data that was extracted were grouped into categories based on notions of similarity, types of IF, and types of mitigation; the categorizations and themes were iteratively refined based on discussions been the authors.

## 3 Results

We identified a total of 1,886 articles through database searches and manually identified one article (see Figure 1). After de-duplication and title and abstract screening, 1,223 papers were excluded, and due to unavailability of full text a further 7 articles were excluded. This resulted in 656 articles for in-depth review, of which 640 were excluded after full-text review. A total of 30 articles (including one identified manually) were studied and analyzed in this review. Table 2 lists and summarizes the 30 articles chosen for inclusion in this scoping review. The articles were numbered sequentially, as shown in Table 2, and this numbering was used to refer to these articles in subsequent tables.

**Table 2:** Articles included in review.

| Article | Description |
| --- | --- |
| Dwork et al.<sup>22</sup> | Original paper introducing the concept of individual fairness and describes a framework for making domain specific similarity metrics. |
| Kusner et al.<sup>41</sup> | Proposes the idea of counterfactual fairness which draws differences in counterfactuals of an individual based on a sensitive attribute. This puts dependence of the goal on the casual structure of the data. |
| Raff et al.<sup>50</sup> | Novel technique using distance to nearest neighbors to inform information gain for fair node splits. Fairness is measured by the cross-validation fold distances of kNNs. |
| Aggarwal et al.<sup>3</sup> | Framework based on counterfactual fairness that uses LIME feature attributions to find an optimal fairness constraint on features used in the model. |
| Garg et al.<sup>28</sup> | Introduces a new fairness metric and three counterfactual based mitigation methods: blindness, counterfactual augmentation, and logit pairing. |
| Sharifi-Malvajerdi et al.<sup>55</sup> | Averages similarity based fairness over many classification models of the data. |
| Binns<sup>13</sup> | Compared theoretical understanding of GF, IF, and their conflicting goals. |
| Bechavod et al.<sup>10</sup> | Uses an auditor for a general adversarial framework of an ambiguous non-parametric similarity between individuals. |
| Hu and Rangwala<sup>34</sup> | Uses two gradient contextual bandit algorithms to compare difference in model behavior for different sensitive feature values to optimize consistency. |
| Kang et al.<sup>37</sup> | Developed methods specific to graph mining based on Dwork et al. and Agarwal et al. |
| Aumuller et al.<sup>7</sup> | Describes a fair clustering algorithm for storing similar individuals in data structures. The current method using LHS had biased probabilities in queries so the paper proposes a uniform ranking system for LHS clusters for consistent probabilities within clusters. |
| Cheng et al.<sup>17</sup> | Research interviewing stakeholders in the medical field to understand their notions of what fairness is in medicine. |
| Fleisher<sup>25</sup> | A criticism of IF and the notion of consistency as a fairness goal. The author argues measuring "similar individuals being treated similarly" is a good way to identify fairness, but constraining the algorithm to that does not ensure generalized fairness. Four arguments are presented against current methods for IF (in terms of consistency). |
| García-Soriano and Bonchi<sup>27</sup> | Introduces a method to achieve IF under GF constraints. Individuals are considered through a learned ranking distribution. |
| Mhasawade and Chunara<sup>46</sup> | Introduced a new form of counterfactual fairness that takes into account not only the individual level causal structures in measuring an individual's fairness, but also the macro causes of sensitive attributes that may affect the outcome differently. |
| Petersen et al.<sup>49</sup> | Post-processing technique proposed where a graph (individuals as nodes and similarity defined by weighted edges) has its adjacency matrix optimized via coordinate descent. |
| Aumüller et al.<sup>8</sup> | An iteration of the previous paper with updates to the same methodology. |
| Chien et al.<sup>18</sup> | Overview of how fairness methods for ML, particularly IF, has room in clinical research. Practical discussion revolves around potential use of reinforcement learning and fairness methods for adaptive clinical trials. |
| Ghadage et al.<sup>31</sup> | Uses a data transformation for fair data representation and uses euclidean distance to measure similar individuals. Multiple sensitive attributes are considered, and the model optimizes the loss using the fairness metric. |
| He et al.<sup>33</sup> | Novel methods regarding applying fairness across continuous spatial data. The authors propose using any loss function in a domain-specific fairness metric to measure disparities between different spatial partitions. Constrained online stochastic learning is used to optimize fairness. |
| Rahman and Purushotham<sup>51</sup> | Novel approach to applying fairness in survival analysis models. The IF application in the paper uses a IF similarity based constraint on the loss function. Models were also shown to be resistant against censoring sensitive features (a potential cause of unfairness). |
| Xie et al.<sup>66</sup> | A visualization framework for fairness methods for graph mining models. The visualizations include known IF methods. |
| Zhang et al.<sup>71</sup> | Proposed an algorithm that can identify discriminatory samples in the data that could cause disparities in predictions. These are used to then retrain a deep learning model for more fair predictions. |
| Biswas and Rajan<sup>14</sup> | A neural network specific solution to IF that partitions input into several strata, prunes neurons from the network, and uses an SMT solver. |
| Caton and Haas<sup>16</sup> | A comprehensive review of fairness methods including GF and IF. The IF methods covered are described briefly and methods are classified into various ML lifecycle groups. |
| Cornacchia et al.<sup>21</sup> | Introduces the concept of "Counterfactual Fair Opportunity" which is an interpretation of Equal Opportunity for counterfactual samples. Additionally, they introduce a new metric, CFlips, that "measures the percentage of counterfactual samples belonging to another demographic group." |
| Ehyaei et al.<sup>23</sup> | Explains a new method of a causal recourse problem called Adversarially Fair Robust Recourse Problem. Under certain conditions, the algorithmic recourse problem can be both individually fair and robust (FARO). |
| Ma et al.<sup>44</sup> | Introduces a learning framework, CLAIRE, that learns a causal model without domain knowledge of the data. Similarity is measured through difference in prediction distributions under different counterfactual values for sensitive attributes. To accomplish this, a VAE model is trained to perform counterfactual data augmentation. |
| Tal<sup>57</sup> | Primarily a discussion on the disparity of definitions and expectations of model outcomes between machine learning models and clinicians. The main conclusion is correlational models tend to model observed outcomes when clinicians expect modeling of counterfactual outcomes. |
| Zhang et al.<sup>70</sup> | A pre-processing mitigation method leveraging matrix estimation. The method handles noisy data with missing values. |

Based on the 30 articles in the review, we identified several themes including philosophical underpinnings of fairness, IF metrics, mitigation methods for achieving IF, implications of achieving IF on GF and vice versa, fairness metrics that combined IF and GF, software for measuring and optimizing IF and applications of IF in healthcare.

### 3.1 Study characteristics

The publication years with the most articles (both with n=7) were 2022 and 2023. Since the seminal article by Dwork et al. was published in 2012, the rate of publication has increased yearly, indicating that this field is still in its infancy, and growth is expected to continue. Similarity in counterfactuals (n=10) was the most common type of similarity. The literature has begun to deviate from a domain-specific distance metric in favor of alternative methods for measuring fairness. Recent publications favor learned distance metrics (n=6) and distance relating to counterfactuals (n=10). Being the original and most intuitive goal for IF, consistency (n=21) was the most prevalent type of IF implied by authors. In-processing methods (n=17) were the popular mitigation type. In our review, we only found a single mitigation method, Petersen et al., that we considered post-processing.^49^

### 3.2 Philosophical corollaries of fairness

IF is motivated by the notion that similar individuals be treated similarly which has been linked to achieving consistency in fairness literature. This notion is linked with Aristotle’s conception of justice as consistency.^53^ It is a desirable aspect of justice that judges render accurate and consistent judgment for every individual and arrive at the same conclusion in identical cases. In the context of algorithms, consistency ensures that an algorithm’s decisions are similar for similar individuals, regardless of group membership. Similarity-based or distance-based measures are commonly used to assess and achieve consistency-based fairness (see next section).

GF is motivated by the notion that groups of individuals should be treated similarly on average when they differ only in protected attributes.^†^ This notion is linked to anti-discrimination laws, which prohibit discrimination against certain groups of people based on protected attributes such as as race, sex, and age. Anti-discrimination in the context of algorithms ensures that an algorithm’s decisions for an underprivileged group are similar on average to decisions for a privileged group. To assess and achieve anti-discrimination fairness, discrimination statistics that measure the average similarity of decisions across groups are used.^58^

In addition to consistency and anti-discrimination, a third concept is counterfactual fairness that ensures an algorithm’s decisions remain consistent across hypothetical scenarios where individuals’ protected attributes are altered.^41^ Typically, causal models that understand how changes in protected attributes affect decisions and other attributes of individuals are used to assess and achieve counterfactual fairness.

### 3.3 Fairness metrics

Measuring IF is typically based on a metric that measures similarity between individuals, and a common way to calculate similarity is by a metric or distance function that defines the distance between individuals as a non-negative real number. Dwork et al. defined an IF metric that assesses the fairness of an algorithm based on if it assigns the same decision to individuals with similar characteristics.^22^ The distance between two individuals, say *a* and *b*, is quantified by a distance measure *d*(*a, b*), and IF is satisfied when

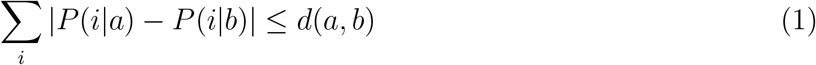

where *P* (*i*|*a*) and *P* (*i*|*b*) are the probabilities of decision *i* for individuals *a* and *b* respectively.^22^ Similarly, Zemel et al. defined an IF metric called the consistency index which assesses the disparity between the decision assigned by an algorithm to an individual and that individual’s k nearest neighbors. The consistency index is expressed as

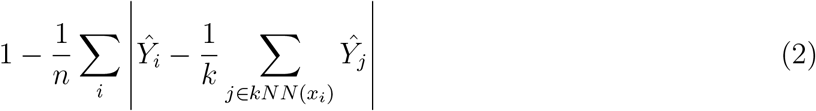

where *n* is the total number of individuals, *Ŷ*_*i*_ is the predicted output for individual *i*, and *x*_*i*_ is the feature vector of individual *i*.

Distance metrics are also used to measure counterfactual fairness Kusner et al. The counter-factual of an individual is a hypothetical scenario in which that individual’s sensitive attributes differ.^41^ The counterfactual fairness metric compares an algorithm’s decision for an individual to their counterfactual. Counterfactual fairness is satisfied when

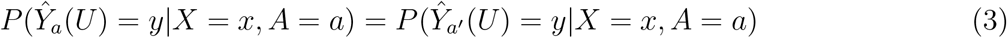

where *Ŷ*_*a*_ and *Ŷ*_*a*′_ are the predicted decisions for an individual and their counterfactual, respectively, defined by sensitive attributes *a, a*′ *∈ A*, latent variables *U*, and feature vector *x ∈ X*. Rather than simply flipping the value of the sensitive attribute(s) to represent the counterfactual, the causal effect of *A → X* is distributed across *X*_*a*_ to derive the features of the counterfactual, *X*_*a*′_. Under this definition, predictions of *P* (*Ŷ*_*a*_) are counterfactually fair if *A* is not a cause of *Ŷ*.

Several types of IF metrics appear in the literature that iterate on the work of Dwork et al., Zemel et al., and Kusner et al. Most simply, generally defined similarity metrics from mathematics such as Euclidean distance, cosine similarity, and Pearson correlation coefficient are widely applicable across various domains and types of data. Domain-specific distance metrics are designed for specific types of data or fields, and they may not be widely applicable outside of their intended domain. For example, Rahman and Purushotham use a derivative of cosine similarity adjusted specifically for hazard-based survival models by Keya et al. Learned distance metrics are derived from the dataset on which they will be applied. Unlike pre-defined distance metrics, learned metrics adapt to the specific characteristics of the dataset (see Table 4). Additionally, counterfactual methods use a variety of unique methods to measure the difference between an individual and their counterfactual. Methods for counterfactual distance vary across each article.

**Table 3:** Philosophical corollaries of fairness. In the Articles column, a key article is highlighted in bold font.

| Philosophical Corollary | Description | Articles |
| --- | --- | --- |
| (Anti-)Discrimination | Ensures that algorithms treat different groups similarly. Discrimination statistics based on demographic parity or equal opportunity, for example, can be used to accomplish this. | 3, 13, 16, 17, 18, 21, 22, 27, 50, 51, 66, **71** |
| Consistency | Ensures that algorithms treat similar individuals similarly, regardless of group membership. Similarity-based or distance-based measures can be used to accomplish this. | 7, 8, **10**, 13, 14, 16, 17, 18, 22, 25, 27, 31, 33, 34, 37, 49, 50, 51, 55, 66, 70 |
| Counterfactual | Ensures that algorithms would have made the same decision for an individual, regardless of their group membership, even if their attributes had been different. Causal models that explain how protected attributes affect decisions can be used to accomplish this. | 3, 13, 16, 18, 21, 23, 28, 41, 44, 46, **57**, 66 |

**Table 4:** Types of IF metrics. In the Articles column, a key article is highlighted in bold font.

| Types of IF metrics | Description | Articles |
| --- | --- | --- |
| Generally defined distance metric | Predefined distance metrics that are widely applicable across various domains and types of data. | **31**, 49, 70 |
| Domain-specific distance metric | Predefined distance metrics that are designed for specific types of data or fields. | 17, 22, 33, 37, **51**, 55 |
| Learned distance metric | Distance metrics that are derived from the dataset on which they will be applied. | 7, 8, 10, 27, **34**, 50 |
| Distance in counterfactuals | Various distance metrics are used to compare an individual to their counterfactuals. | 3, 14, 21, 23, 28, 41, **44**, 46, 57, 71 |

**Table 5:** Types of mitigation methods. In the Articles column, a key article is highlighted in bold font.

| Type of Mitigation | Description | Articles |
| --- | --- | --- |
| Pre-processing | Adjust, transform, reweight, or augment data to ensure balanced representation and remove discrimination. | 3, 10, 14, 23, 28, 31, 41, 44, 46, 57, **70**, 71 |
| In-processing | Modify the training of the algorithm to incorporate fairness constraints or objectives directly. | 7, 8, 10, 14, 22, 23, 27, 28, 31, 33, 34, 37, 44, 50, 51, **55**, 71 |
| Post-processing | Adjust or transform the decisions or outputs of an algorithm subsequent to its training. | **49** |

### 3.4 Mitigation methods

In the context of creating fair algorithms, pre-processing, in-processing, and post-processing are three categories of methods to mitigate bias. Pre-processing methods adjust or transform the data to ensure balanced representation and remove discrimination. Examples include resampling (adjusting the data to balance the representation of different groups^3^), reweighting (assigning different weights to samples to counteract imbalances^16^), and removing protected attributes (removing features like race, gender, or age that are protected and could lead to biased decisions^28^). In-processing methods modify the training of the algorithm to incorporate fairness constraints or objectives directly. Examples include regularization techniques (adding a fairness constraint or regularization term to the learning objective^37^) and adversarial debiasing (using adversarial networks to learn representations that do not contain biased information about protected attributes^14^). Post-processing methods adjust or transform the decisions or outputs of an algorithm subsequent to its training. Examples include calibration (adjusting predicted probabilities of decisions to reflect the true likelihoods of those decisions accurately) and threshold adjustments (changing decision thresholds for different groups to balance performance metrics).^16^

Although pre-processing and in-processing techniques were frequently employed in the articles we reviewed, post-processing to reduce bias was only described in one article.^49^

### 3.5 The relationship between the two kinds of fairness

Several GF metrics are incompatible in that fairness cannot be achieved simultaneously on those metrics. The incompatibility of IF and GF metrics has received less attention. GF does not imply IF, and IF implies GF if the Wasserstein distance (distance between probability distributions) is small, i.e., the distributions of similar individuals are relatively uniform across groups, which is uncommon in practice. ^22, 72^

Binns discusses the trade-offs that arise when one type of fairness is preferred over another. When ignoring IF in favor of GF, algorithms may make different decisions for identical individuals. Furthermore, emphasizing IF alone can lead to significant differences in group decisions.^13^ According to Fleisher, optimizing IF alone does not guarantee GF. An algorithm that assigns a negative decision to every individual, for example, will satisfy IF but not GF.^25^

### 3.6 Combining GF and IF metrics

Speicher et al. describe an index for overall unfairness that measures model inequality by assessing each individual’s level of favorable treatment. Furthermore, when the data is divided into distinct groups, they demonstrate that this metric can be rewritten as the sum of two new measures known as between-group and within-group unfairness, which correspond to the group-level and individual-level bias indicators discussed here. As a result, the overall unfairness index evaluates discrimination on both levels. The authors then use this decomposed expression to show the potential imbalance in group and individual fairness.^56^

Furthermore, Friedler et al. explain this incompatibility between groups and individuals by first defining two opposing fairness perspectives, which they call WYSIWYG (what you see is what you get) and WAE (we’re all equal). They also define three information spaces: the unobserved, the observed, and the decision spaces. The unobserved and observed information spaces are the same from a WYSIWYG perspective. WAE, on the other hand, assumes that there are no discriminatory differences between groups in the unobserved space, but bias is introduced in the observed space beyond an individual’s control. Whereas the WYSIWYG view has little distortion between the two spaces, i.e. the distance between individuals in the observed space and the decision space is generally the same (maintains individual fairness), WAE reduces the distance between groups in the decision space (upholds group fairness).^26^

Zemel et al. describe a method that achieves both GF and IF. The method works by finding a good representation of the data with two competing goals: to encode the data as well as possible, while simultaneously obfuscating any information about membership in the protected group.^69^

### 3.7 Software

Several articles from the review provide software implementations seen in Table 6. Additional IF concepts are implemented in open-source software. A prominent Python package that focuses on IF, *inFairness*,^67^ was developed by IBM and is designed for PyTorch models. It includes various metrics, metric learning algorithms, in-processing mitigations, and an implementation of Petersen et al.’s post-processing method. Another popular software developed by IBM including IF concepts is *AI Fairness 360*,^11^ which is implemented in both Python and R. The Python package *FairPy* includes other IF methods such as fair rankings similar to Aumuller et al. and García-Soriano and Bonchi ^7, 27^

**Table 6:**
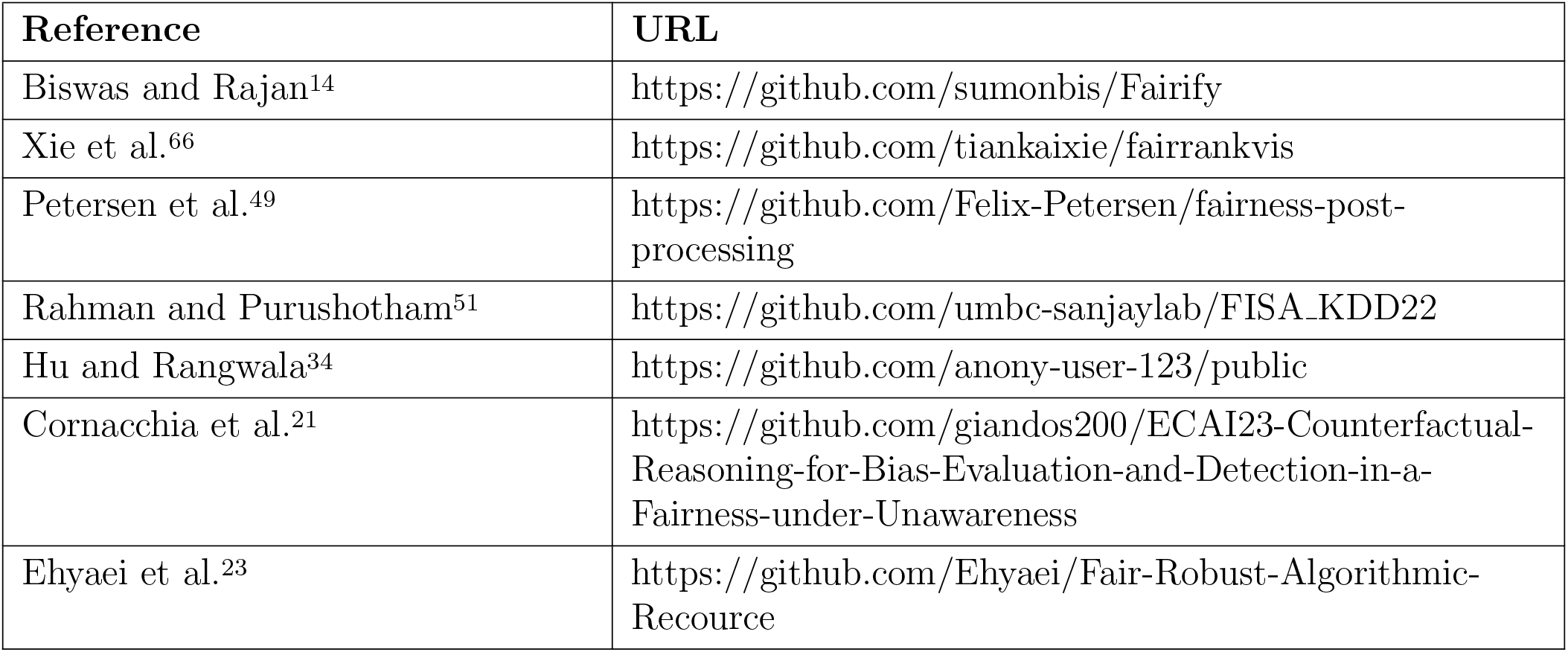
Available software related to IF.

### 3.8 Applications in healthcare

Applications of IF in healthcare were discussed in Cheng et al., Chien et al., Rahman and Purushotham, and Tal.^17, 18, 51, 57^ Rahman and Purushotham describe an IF method for survival analysis to address the problem of censoring in clinical trials, particularly in underprivileged groups.^54^ The authors demonstrated that their IF-based deep survival algorithms reduced unfairness in censoring significantly.

Cheng et al. created a framework for conducting interviews with stakeholders to better understand their interpretations and notions of fairness in clinical predictive systems. Twelve participants were polled, and many of them were skeptical of IF. For example, one participant remarked, “I think it’s tricky to compare things this way,… It’s hard to say.” Although more participants favored GF, they disagreed on which GF measures were appropriate.^17^

Chien et al. suggest that the traditional fixed-clinical trial method prevents beneficial modifications after trials begin, and AI methods can be employed to make trials fairer. According to the authors, optimizing for GF is less useful than optimizing for IF or CF for the problem of fairness in clinical trials, despite the advantage of GF methods being task-agnostic and less complex.^18^

Tal argues that an important cause of bias in healthcare algorithms is due to conflicting notions of problem definitions. For example, a statistical notion of bias and accuracy would claim the two are orthogonal, allowing a model to be both biased and accurate. On the other hand, a clinician would argue that bias and accuracy are contradictory and cannot co-occur. Target specification bias, a particular case of this divergence in definitions, occurs when the notions of the decision variable by analysts and clinicians differ.^43, 57^ This occurs because a clinician expects to predict a decision for a patient if they were treated differently all else being equal (counterfactual), whereas most models predict similarly observed individuals with measured decisions. This issue is closely related to IF, implying that CF is a more accurate representation of the problem from the standpoint of a clinician.

## 4 Discussion

Given the relatively small number of articles we found (30 articles from 2013 to 2023), the first implication of our findings is that the definition, use, and study of IF remain in their infancy, especially in healthcare. However, since the seminal article on IF was published in 2012, the rate of IF article publication has steadily increased, indicating that this field is likely to grow in the future. Only four articles described the use of IF in healthcare, despite evidence that there is intense interest in measuring and mitigating bias in clinical risk calculators based on race, differential laboratory test reference ranges are recommended based on race, and differential therapy is recommended based on race.^63^ This is most likely due to the infancy of the field of IF in general.

There is mounting evidence that due to the limitations of GF, alternative approaches to fairness, such as IF and CF, are needed. One limitation of GF is that it may mask individual differences within a group, whereas IF is more flexible and adaptable and can take individual features other than protected attributes into account. Second, GF may disregard relevant features that are not protected attributes, whereas IF may lead to more accurate and fair outcomes by considering all relevant features. Third, defining and measuring GF can be challenging when dealing with multiple groups with overlapping memberships or complex relationships, while IF is not limited in such circumstances. While IF offers several advantages over GF, it also has limitations that need to be considered. One limitation of IF is that determining what constitutes “similar individuals” can be complex and subjective. Different contexts and tasks may require different definitions of similarity, making it challenging to achieve universal applicability. Second, IF methods that rely on learning similarity metrics from data are susceptible to encoding existing biases present in the data, which can perpetuate existing inequities.

While GF ensures balance across groups, it can mask individual discrimination. Conversely, IF may be susceptible to biased definitions of similarity. Combining both notions of fairness may offer more robust solutions. Several articles, including Speicher et al.and Zemel et al. described methods that combine the opposing goals of GF and IF.^56, 69^

Several articles discussed CF, which focuses on whether the outcome for an individual would be different if their protected attributes were different, holding all other features constant. This “what-if” scenario relies heavily on causal models, which provide a framework for understanding the relationships between features and outcomes. Causal models are crucial for CF, as they determine whether an individual’s outcome is truly influenced by their protected attributes or other relevant features. By incorporating causal models, CF brings fairness methods closer to expectations of fairness by clinicians, for example, addressing target specification bias,^57^ or designing adaptive clinical trials.^18^

Our study had some limitations. By limiting our search to three databases, it is possible that articles relevant to this topic were not found and included in this review. It is also possible that relevant articles that used keywords other than those that were used were missed in our search. It is clear from our search results that the number of articles in IF has increased steadily over the last decade. However, we did not investigate how IF metrics and definitions have evolved over this period of time.

## 5 Conclusion

This scoping review explored the breadth of algorithmic IF metrics and methods developed to achieve IF. The articles that explored this topic showed that the definition, use, and study of IF remain in their infancy, especially in healthcare. Future research is needed in the application and evaluation of IF for healthcare.

## Data Availability

All data produced in the present work are contained in the manuscript.

## 6 Acknowledgements

Research reported in this publication was supported by the National Institutes of Health under award number T15 LM007059 from the National Library of Medicine and under award number UL1 TR001857 from the National Center for Advancing Translational Sciences. It was also supported by a School of Computing and Information Predoctoral Fellowship to JWA.

## Glossary

Fairness metric: A mathematical definition of “fairness” that is measurable.
Fairness constraint: Applying a constraint to an algorithm to ensure one or more definitions of fairness are satisfied.
Group Fairness: An algorithm is fair if some statistic of the algorithm is equal across protected groups.
Individual Fairness: An algorithm is fair if it gives similar predictions to similar individuals.
Counterfactual: The unobservable case where an individual belonging to some “treatment” group, *a*, were to belong to an alternative group, *a*′.
Counterfactual Fairness: An algorithm is fair if gives similar predictions for individuals and their counterfactuals.
Target specification bias: A discrepancy in the operationalization and understanding of a target variable between clinicians and researchers.
Consistency: A philosophical principle of fairness of “similar individuals being treated similarly.”
Discrimination: A philosophical principle of fairness that the performance of a predictive model should be similar or equal across protected groups.
Pre-processing: Adjust, transform, reweight, or augment data to ensure balanced representation and remove discrimination.
In-processing: Modify the training of the algorithm to incorporate fairness constraints or objectives directly.
Post-processing: Adjust or transform the decisions or outputs of an algorithm subsequent to its training.
Individual justice: The idea that individuals should be assessed on their own qualities, circumstances, and attributes, not based on generalizations about groups of which they happen to be a member.

## Footnotes

† In the biomedical literature, models are frequently referred to as algorithms. In this paper, we use the terms algorithm and model interchangeably and preferably use the term algorithm.

† In this paper, we use the terms protected and sensitive interchangeably and preferably use the term protected.

